# Investigating biomarkers of exposure to jet aircraft oil fumes using mass spectrometry

**DOI:** 10.1101/2025.04.17.25326021

**Authors:** Clement E. Furlong, Rebecca Richter, Judit Marsillach, Alex Zelter, Matthew McDonald, Allan Rettie, Oksana Lockridge, Rachel Lundeen, Dale Whittington

## Abstract

Most commercial passenger jet aircraft use compressed engine air after cooling as a source for ventilation and cabin pressurization onboard. This “bleed air” design means that engine oil and/or hydraulic fluid can contaminate the ventilation supply air during otherwise normal flights, exposing onboard crewmembers and passengers to the fumes. The oils and hydraulic fluids contain a complex mixture of triaryl phosphates (TAPs) and decomposition products. Although the health and flight safety consequences of inhaling these fumes have been widely documented, measures of onboard inhalation exposure have been lacking. An approach is presented for documenting exposure to engine oil fumes by using high-resolution mass spectrometry (MS) to monitor and quantify post-translational modifications of subjects’ butyrylcholinesterase (BChE) that are consistent with exposure to the engine oil TAPs. We hypothesized that plasma from exposed individuals would show modifications or adducts on the active site serine (Ser198) of BChE. Plasma BChE from 81 exposed subjects was purified to near homogeneity and concentrated using antibodies immobilized on paramagnetic beads. The purified BChE was eluted at low pH, neutralized, digested with trypsin, and analyzed by liquid chromatography (LC)-MS. In subjects reporting onboard oil fume exposures, the most consistent adduct modifying the Ser198-containing tryptic peptide had a mass value of +154.0031 Da. The normalized peak area (NPA) of the +154Da modification was determined by comparing the relative MS1 intensities of the +154Da-modified Ser198 containing peptide to the total observable peptides containing the active site, including missed cleavages. Notably, adducts from *in vitro* exposures of bioactivated TAPs to purified BChE conducted in this study (i.e., +80Da, +156Da, +170Da, and +186Da) as well as adducts reported in other earlier in vitro studies (i.e., +65Da, +80Da, +91Da, +107Da, +165Da, +180Da, +181Da, and +277Da) were not detected in exposed subjects. Of the 81 subjects in this study, the average NPA of +154Da-Ser198 resulted from fume event exposures that pre-dated 2013 (N=59); range = 0.46%-17.8%, *with X̅* =4.84% which was 9.7 times higher than control subjects (*X̅*= 0.50%) These data are uncorrected for the time lag between the reported exposure and the blood draw. Samples from the remaining 16 subjects with exposures from 2016-2024 showed only the 154Da modification at background levels (0.24%-1.13%; *X̅*=0.55%), as confirmed in control plasma samples from individuals who had not flown in at least three months. The observed reduction in the 154Da adduct over time in exposed individuals is likely a function of the change in the formulation of the OP blends added to engine oils during the course of the study. Further investigation into other protein biomarkers and adducts correlated with exposure to the current oil additives and hydraulic fluid fumes on aircraft is warranted. The most satisfactory solution would be to eliminate the exposure hazard by implementing bleed-free systems or, at a minimum, to develop less toxic oil formulations, suitable bleed air filters, and modified designs.

## Introduction

Tricresyl phosphates (TCPs) and other triaryl phosphates (TAPs), a subclass of aromatic organophosphates (OPs), are added to aircraft engine oils to reduce wear and improve thermal stability in the engines. In contrast, aircraft hydraulic fluid composition is dominated by alternate TAPs and alkyl phosphates. Importantly, the chemistry of these additives is significant beyond aircraft engine performance. The design of almost all aircraft permits heated oils and hydraulic fluids to infiltrate and contaminate the ventilation air supplied to the flight deck and cabin. Outside air used to ventilate and pressurize the cabin is extracted from an engine compression system, resulting in contamination of the air supply with engine oil fumes. This can happen at low levels during routine operations (often during engine power setting changes) and far less frequently when an engine seal is defective or fails. Despite the documented health and flight safety consequences, measurements of onboard inhalation exposure to these fumes have been lacking.

Historically, inhibition of the enzymatic activity of plasma cholinesterase – specifically butyrylcholinesterase (BChE) – has served as a standard indicator of exposure to various neurotoxic OPs (Wilson and Henderson, 2007). In this context, modifications to BChE induced by TAPs provide a surrogate measure for assessing exposure to engine oil fumes. However, while this biochemical marker offers insights, the broader health implications of inhalation and exposure extend well beyond enzyme inhibition. The neurotoxic properties of specific OP compounds have been exploited in insecticides and nerve agents, and more recently, they have been used as anti-wear agents in aircraft engine oils.

Tricresyl phosphates – especially the isomers with an ortho methyl group – have been recognized as potent neurotoxins since the 1930s (Henschler, 1058), prompting early concern by the United States military in the early 1950s for its use as a major component in anti-wear formulations (Kitzes, 1956; Loomis & Krop, 1955 and Reddall, 1955). In fact, Reddall proposed solutions for eliminating the OPs from cabin air, ranging from the use of special filters to the use of compressors independent from the engines for conditioning the cabin air. Today, only the Boeing 787 aircraft uses dedicated electrically driven compressors for ventilation which avoids engine oil contamination of the air supply. Airline fume and smoke data that US airlines are required to report to the Federal Aviation Administration (FAA) authority reflect these design differences. Over a recent four-year period, engine oil fumes were the most frequently reported incident. As expected, though, US airlines did not report any smoke/fume events on B787 aircraft during those four years (Anderson, 2025). On the other hand, individuals exposed to oil or hydraulic fume events from the aircraft using bleed air for ventilation supply may suffer significant health impacts. In such cases, exposure to oil or hydraulic fumes has been associated with symptoms severe enough to result in loss of certification to fly for pilots and an inability to return to work for cabin crew members (Michaelis, 2017).

Motivated by these concerns, the primary objective of this research was to develop one or more biomarkers to document exposure to TAPs found in aviation engine oil fumes. By developing and validating mass spectrometry methods that laboratories worldwide can adopt, this approach aims to establish robust protocols for monitoring exposure to these harmful chemicals. This work aims to improve our capacity to document such exposures and provide insights that can guide strategies to protect flight crews and passengers from the adverse health effects associated with contaminated cabin air.

## Materials and Methods

### Materials

Dynabeads™ Antibody Coupling Kit M-270 14311D, DynaMag™-2 Magnet, anti-Butyrylcholinesterase Monoclonal Antibody (3E8), Eppendorf Protein LoBind^®^ microtubes, Greiner Bio-One #655101 96-well flat bottom microplates for visible light, Water Optima™ W6-4, acetonitrile (ACN) Optima A955-4, formic acid LC/MS grade Optima™ A117-50, PBS, iodoacetamide No-Weigh™ Format, Pierce™, Dithiothreitol (DTT) No-Weigh™ Format, Zeba™ Spin Desalting Columns 0.5 mL 7K MWCO were purchased from Fisher Scientific (Hanover Park, IL). Mass spectrometry grade acetic acid, Tween 20, Butyrylthiocholine iodide (BTC), 5,5′-dithiobis-(2-nitrobenzoic acid (DTNB), were from Sigma-Aldrich (St. Louis, MO). Sequencing Grade Modified Trypsin was from Promega (Madison, WI). Human Liver Microsomes (HLMs) were from SEKISUI XenoTech (Kansas City, KS). NADPH was from Research Products International (Mt. Prospect, IL). Durad 125, Durad 150, and Syn-O-Add 8484® were acquired from commercial sources. Mono- and di-hydroxy methyl T*m*CP were a gift from Dr. Matt McDonald. BChE collected and purified from a pool of approximately 8,000 plasma samples (pooled BChE or pBChE) served as a “BChE standard” and was a gift from Dr. Oksana Lockridge.

### Description of subjects

The present study was approved by the University of Washington human subjects review process and all subjects provided consent or assent forms. We invited airline crew members who had documented an onboard exposure to oil fumes to send blood samples to our laboratory for analysis. Outreach was largely conducted by way of safety department representatives of airline crewmember unions, including those in the US, UK, Canada, Australia, France and Germany. Samples from individuals who had reported onboard conditions consistent with exposure to oil fumes, often with attendant symptoms, voluntarily shipped blood samples to our laboratory with a consent form and brief questionnaire regarding the onboard conditions. Control blood samples, also with a consent form, were provided by individuals who had not flown for at least three months.

### Blood Draws

Venous blood samples were drawn into lithium heparin anti-coagulant tubes (Tube BD 367886). EDTA tubes are also acceptable for the protocols. The lithium heparin tubes were chosen because at the onset of this study we did not know whether the HDL-associated enzyme paraoxonase 1 (PON1), that protects against the toxic metabolites of two organophosphorus insecticides chlorpyrifos and diazinon, would also be protective against TAP exposures. EDTA is a potent inhibitor of PON1 activity (Ortigoza-Ferado, 1984). Varying volumes of venous blood (6-20 mL) were drawn at different phases of the study. Our current protocol involves drawing two 10 mL samples of blood. To simplify and standardize the sample processing and shipping, blood samples are collected into two 10 mL blood draw tubes which are then inserted into 50 mL Falcon™ tubes to contain any leakage that may occur during shipment. The tubes were shipped to our laboratory overnight on gel ice without freezing which would lyse the red cells (RBCs). When the samples arrived, the blood cells were sedimented by centrifugation in a Beckman J6-B swinging bucket centrifuge with a JS-3.0 rotor for 5 min at 2500 RPM (1780 x g) or equivalent procedure. The plasma was carefully removed, and the two fractions (plasma/RBCs) were aliquoted into 1.5 mL aliquots in labeled 2 mL microfuge tubes then frozen at -80°C.

### Preparation of Magnetic Beads for Immunopurification of Human Butyrylcholinesterase (BChE) from plasma

Monoclonal mouse anti-human BChE antibodies were attached to Epoxy Dynabeads™ according to the manufacturer’s instructions with supplied buffers in Dynabeads KIT 14311D. Eppendorf Protein LoBind® microtubes (0.5, 1.5, or 2.0 mL) were used with all magnetic bead protocols. One hundred µg of anti-human BChE antibody were attached to 10 mg of Epoxy Dynabeads™ in one 1.5 mL LoBind® microtube. First, 10 mg of Epoxy beads were hydrated in 1 mL of buffer C1, rotated at RT for 15 minutes. The C1 buffer was removed after immobilization of the beads by magnet, and 400 µl of fresh buffer C1 were added. BChE Antibody, 100 µg in 100 µL were added, followed by 500 µl of buffer C2 for a final volume of 1 mL in one 1.5 mL LoBind® microtube with overnight end over end rotation at 37°C. Following incubation, the supernatant was removed using a DynaMag™-2 Magnet to immobilize the beads with bound antibody. A short, 2 second centrifugation was used as needed to pellet tube contents away from the top of the tubes before applying the tube to the magnet while working with the beads. The beads were then washed sequentially with 1 mL each of the provided proprietary Dynabead kit buffers: HB first, LB second, and finally SB buffer using the magnet to separate the beads from the supernatants at each step. A final wash using 1 mL of buffer SB was performed by rotating the tube for 15 minutes at room temperature. The final supernatant was removed, and 1 mL of fresh SB buffer was added for storage of the antibody-conjugated beads at 4°C. The resulting magnetic beads had 100 micrograms of anti-BChE antibody attached to 10 mg of epoxy beads in a volume of 1 mL SB buffer. The antibody coupled beads were stored at 4°C until use. Just prior to use, 1 mL of beads were washed 3 times with 1 mL of PBS using the magnet to immobilize the beads while removing the wash solution. The antibody-bead conjugate was stored in the final 1 mL wash at 4^°^C until used.

### Immunocapture of Human BChE from plasma

BChE from plasma samples was purified as follows: plasma samples were thawed at room temperature, mixed well, and centrifuged at 14,000 rpm (20817 x g) in a microcentrifuge for 5 minutes to remove any precipitate. Five hundred µL samples were transferred to fresh 1.5 mL microfuge tubes. The samples were diluted with 500 µL of PBS and mixed well. 10 µL of each initial diluted plasma sample were used to quantify the efficiency of the bead capture protocol with the BChE activity assay (usually 80-95% yield). Any leftover undiluted plasma from each thawed sample was refrozen. Freshly washed, well suspended 100 µL aliquots of magnetic bead-antibody complex in PBS were transferred to 1.5 mL Protein LoBind® microtubes and placed into the magnet to immobilize the beads to the side of the tube. In all cases with magnetic beads prior to placing the tubes against the magnet, a brief 1-second microfuge spin was used to ensure all the tube contents were away from the top of the tube. The PBS was removed, leaving the beads (∼1 mg) complexed to anti-BChE antibody in the LoBind® microtubes. The 1.0 mL of diluted plasma sample was added to each tube of beads and mixed well. The beads and sample were rotated end-over-end at room temperature for two hours. Next, the sample was transferred to the magnet to capture the bead-bound BChE from the diluted plasma. The removed depleted plasma samples were used to assay the percentage of BChE removed by the immunobead protocol. BChE activity in both the diluted plasma sample aliquots taken before and after exposure to the antibody-conjugated immunomagnetic beads was measured with the BChE assay (Ellman 1961) adapted for the plate reader. In all steps, great care was used to separate all the liquid phase supernatants from the magnetic beads in the Protein LoBind®microtubes. The leftover diluted BChE depleted plasma samples removed following the two-hour exposure to the beads were refrozen at -80°C for possible future use in identifying other biomarker protein targets.

The 100 µL of magnetic beads with bound BChE were washed twice with 1 mL of PBS containing 0.5% fresh Tween 20, discarding both washes and leaving the washed beads. The beads were then transferred to a fresh 1.5 mL LoBind® tube using 1 mL PBS without Tween 20 and washed once more with 1 mL PBS to remove any remaining Tween detergent. Following the second PBS wash, the beads were washed once with 1 mL of 10 mM ammonium bicarbonate pH 8.0 (AMBIC). After removing the 1 mL wash, 0.5 mL of the same buffer was used to transfer the beads to a fresh 0.5 mL LoBind® microtube. After immobilizing the beads and removing the 0.5 mL AMBIC wash, 0.5 mL of HPLC grade water was added to wash the beads just prior to elution, then removed after immobilizing the beads with the magnet. The water washed beads were eluted using 70 µL of freshly prepared 6% MS grade acetic acid in HPLC grade water. The tubes containing the beads were mixed vigorously for 3 minutes with the acetic acid eluant. The beads were immobilized with the magnet and the 70 µL of eluted BChE was carefully moved to a fresh 0.5 mL LoBind® tube. The eluted BChE was measured for protein content using a Nanodrop spectrophotometer. If it is useful to retain BChE activity, high pH can be used to release the BChE from the beads using 100 µL 1 M NH_4_OH and neutralizing the extract with 54 µL of MS grade acetic acid after transferring released BChE to a LoBind® tube.

### Measurement of BChE activity

BChE activity was determined by a kinetic modification of the Ellman procedure (Ellman et al. 1961) adapted for continuous activity monitoring in 96 well visible microplates with the plate reader. Kinetic data was acquired at 405 nm for 4 min at RT using SoftMax Pro software, with path length correction. Only linear initial reaction rates (< 4 min or linear portion of the rate curves) were used for analyses. BChE activity was measured as follows: One hundred µL of appropriately diluted BChE samples were deposited in microwells in triplicate. Just before use, a 2X concentrated substrate solution of butyrylthiocholine iodide, BTC, (Sigma B 3252) and DTNB (5,5’-dithio-bis-nitrobenzoic acid, Sigma D-8130) was prepared as follows. A solution of DTNB was prepared as a 10.3 mM stock solution in 100 mM sodium phosphate pH 7.0. The final 2X substrate solution contained 0.64 mM DTNB and 2.0 mM BTC in 100 mM sodium phosphate pH 8.0. The reaction was initiated by the addition of 100 µL of 2X substrate solution to the BChE samples with a multichannel pipette.

### Small Scale Microsomal Bioactivation of TAPs for kinetic studies

To examine the inhibition of BChE by bioactivated TAPs three TAPs (T*o*CP, T*p*CP, T*m*CP) and three TAP formulations added to the range of aviation engine oils (Syn-O-Ad 8484®, Durad 150, and Durad 125), and trimethylolpropane phosphate (a possible high temperature reaction product of TCP with the trimetholyl ester base stock of one type of commercial oil) were bioactivated as follows. The six organophosphates were diluted into acetonitrile (ACN) at 2.5 mg/mL. They were further diluted just prior to use at 40 µg/mL in 50 mM sodium phosphate pH 7.4, prior to further diluting to the noted concentrations in a 100 µL final volume in microplate wells in triplicate. Five OPs (except T*o*CP) were diluted to six final concentrations from the 40 µg/ml freshly diluted stock solution. The final concentrations for five of the OPs were, 10, 7.5, 5.0, 2.5 1.0, and 0.5 µg/mL in a 100 µL final volume. The T*o*CP was further diluted to 0.25, 0.1, 0.025, 0.01, 0.005 and 0.002 µg/mL in the 100 µL final volume. The 100 µl final volumes of bioactivation mixtures contained either OPs alone, OPs with 50 µg/ml HLMs in 50 mM sodium phosphate pH 7.4 (no NADPH), or OPs with 50µg/ml HLMs and NADPH at 1 mM final concentration (complete assay). Bioactivation proceeded for 25 minutes at 25°C in the microplate wells. Following bioactivation, 10 µL of BChE (0.19 µg) were added to each microwell with a multichannel pipettor (Matrix Equalizer ThermoFisher), mixed with the pipettor, and incubated for 30 minutes at 25°C. BChE substrate buffer (100 µL) containing 2 mM BTC and 0.64 mM DTNB in 100 mM sodium phosphate pH 8.0, was added to all microwells to initiate the assay which was monitored over 4 minutes at 405 nm in the microplate reader. Plate reader readings in mOD/min at A_405_ were divided by the path length of each well.

### In vitro OP exposure of purified BChE for LC-MS characterization

The small quantities of bioactivated metabolites required to inhibit pBChE activity for kinetic studies were insufficient for generating the levels of adducted BChE needed for MS analysis. It was necessary to increase the scale of the generation of bioactivated inhibitors. To achieve this goal, pBChE was inhibited as follows:10 mL of 25 µg/mL of OP (D125, D150, T*o*CP, tri–butyl phosphate, or T*m*CP) were placed in a Falcon® 14 mL Round Bottom snap-cap tube (Falcon 352059). HLM’s were added to 150 µg/ml, followed by the addition of 2 mM final NADPH. The tube(s) were rotated end over end at RT for 1 hour. These bioactivated mixtures were then filtered through an Amicon® Ultra-15 Centrifugal Filter Unit (10 kDa MWCO) at 2500 rpm for ≈1.5 hours. The filtrate was then moved to a new snap-cap tube. Then, 4.4 µg of pBChE was added to the 9 mL of filtered bioactivated TAP and rotated at RT for 1 hour while the extent of the inhibition was monitored by BChE assay using 20 µL of the reaction volume. Each BChE sample (4.4 µg) was inhibited to up to 90% as determined by activity analysis. The inhibited pBChE was removed from the incubation mix by adding anti-BChE antibody conjugated paramagnetic beads as follows. New beads (200 µL per 4.4 µg inhibited BChE sample) were washed with PBS by transferring 200 µL of beads to a 1.5 mL LoBind® tube. They were washed 3 times with 1 mL of PBS by magnetic immobilization of the conjugated beads followed by removal of the PBS liquid. The antibody conjugated beads were finally resuspended in 100 µL of PBS and transferred to each of the 9 mL samples of inhibited BChE. The antibody-conjugated beads and bioactivated TAP/ BChE solutions were again rotated at RT for one hour. The percentage capture of inhibited BChE was determined by measuring the inhibited BChE activity remaining over time by sampling 20 µL of the supernatant solution after immobilizing the bead/BChE complexes with the magnet. The measurements indicated that the beads bound up to 90% of the residual BChE activity in one hour. The beads with bound BChE were immobilized with the magnet and the supernatant solution was removed. Each 200 µL of beads were washed in 1.5 mL LoBind® tubes twice with 1 ml of PBS, once with 10 mM AMBIC pH 8.0, and finally with 1 mL of ultrapure water. The 200 µL of BChE-bound beads with water wash removed were eluted with 160 µL of 6% MS grade acetic acid by mixing well for 3 minutes. The stripped beads were again immobilized with the magnet and the acetic acid eluate was transferred to a 0.5 mL LoBind® tube for trypsin digestion as noted above for 2.5h.

### Trypsin Digestion of Eluted plasma BChE

The acetic acid eluted BChE was brought to a final volume of 100 µL with ultrapure water and desalted into 100 mM MS grade ammonium bicarbonate pH 8.0 (AMBIC) using 0.5 mL 7,000 MWCO Zeba™ spin desalting columns that had been equilibrated 3 times with 0.3 mL of AMBIC. The 100 µL eluted samples were carefully added and desalted by centrifuging at 1500 × g for 2 min to collect the desalted sample. The samples were heated to 90°C for 5 minutes, then cooled to room temperature. DTT was added to 5 mM final by adding 1 μL of 500 mM Pierce No-Weigh DTT in water. Samples were then incubated at 50° C for 30 minutes and cooled to room temperature. Pierce No-Weigh iodoacetamide was freshly resuspended to 375 mM with132 µL of 100 mM AMBIC and 4 μL were added to each sample and mixed for a 15 mM final concentration. The sample was incubated at room temperature in the dark for 30 minutes. Promega Sequencing Grade Modified Trypsin (1 µg) was added to each sample, mixed well, and incubated at 37° C for 2.5 h. To stop the digestion, 10% MS grade formic acid was added to 0.5% final concentration. Samples were stored frozen at -20°C prior to MS analysis.

### Mass spec analysis of samples

Digested tryptic peptides were injected onto a Waters Acquity I-Class Ultra High-Pressure Liquid Chromatography system (UPLC) using a Waters Acquity UPLC Peptide CSH C18 column (100mm length x 1mm ID, 1.7µm, 70°C). Peptides were separated over a 50-minute gradient method at a flow rate of 0.1mL/min with mobile phase consisting of 100% LC-MS grade water with 0.1% formic acid (A in water) and 100% LC-MS grade acetonitrile with 0.1% formic acid (B), starting at 1%B for 0 to 2 min, increasing to 12% B from 2 to 5 min, then ramping from 12% to 40% B over 30 min. The gradient then switched to 100% B for 2 min and held at 100% for another 2 min before ramping back and equilibrating to starting conditions of 1% B from 40 to 50 min. The UPLC was coupled to a Thermo Ascend Orbitrap Tribrid high resolution MS equipped with a heated electrospray ionization source. All LC-MS analyses were carried out in positive ion mode at a spray voltage of 3500 V with the ion transfer tube set at 275°C and vaporizer temperature at 30°C; additional ion source parameters were as follows: sheath gas 12, aux gas 2, sweep gas 2.8. For peptide mapping and modification analysis, the MS^1^ scan resolution was set to 120,000 in the Orbitrap, range was 300-2000 *m/z*, the normalized AGC target was set to 100%, RF Lens 60% and the maximum injection time was 251 msec. All MS1 data were acquired in profile mode. A filter for peptide monoisotopic peak determination was used along with an 8 sec dynamic exclusion duration time. MS/MS data were collected in data-dependent acquisition with a cycle time of two seconds between master scans. Data-dependent MS2 (ddMS^2^) was conducted in the ion trap at rapid scan rate and was acquired, first using HCD and then CID, for one scan. Settings for both ddM2 scans used the quadrupole isolation window of 2 *m/z*, auto scan range, standard AGC target, and auto maximum inject time. MS2 data acquired in centroid mode. For HCD activation, a normalized collision energy of 30% was used. For CID activation, a collision energy of 35% was used with 10 msec activation time.

### Database searching for variable PTMs and spectral library generation

Database searching and validation were conducted using the open-source software FragPipe (v22.) Raw data were first converted to mzML (MSconvert) and searched using MSFragger against a FASTA protein database consisting of human proteins (20417 total protein sequences, accessed November 2023) and concatenated with a set of reversed sequences. Initial searches were done using open or common mass offset workflows to help elucidate mass adducts. MSFragger search parameters included trypsin enzyme specificity, up to three missed cleavage sites, and variable clipping of N-terminal methionine. Carbamidomethylation of cysteine residues (+57.0125Da) and oxidation of methionine residues (+15.9949 Da) were both set as variable modifications. Further searches in MSFragger utilized closed searching parameters and were evaluated as a function of several factors: enzyme specificity (tryptic vs. no enzyme), number of allowed non-tryptic termini and number of missed cleavage sites as well as variation of fixed and variable post-translational modifications (PTMs). Precursor and fragment mass tolerance was set to 20ppm. The optimized parameters described here had the topmost peptide- and PTM-identifications for in vitro pBChE modifications as well as subject data: tryptic enzyme specificity, up to 3 missed cleavages, clipping of N-terminal methionine (variable), carbamidomethylation of cysteine residues (+57.0125Da, fixed) and the following variable modifications on serine residues: phosphorylation (+79.96633Da), +154 modification (+154.0031Da), +156 modification (+155.99763Da), cresyl phosphate (170.01328), +186 modification (+186.0082Da), and +198 modification (+198.04459Da). For peptide and protein identification, FragPipe .pepxml files were submitted to PeptideProphet and ProteinProphet, with a false discovery rate set to less than 1% based on the decoy database search. Output as .pepxml files were compiled within Skyline to generate spectral libraries for peptides at greater than 90% probability.

### Label-free peptide quantification using MS1 peak area filtering

All LC-MS data acquired from subjects and in vitro studies were analyzed in Skyline. Skyline template files containing all possible combinations of PTMs on BChE tryptic peptides were generated. Spectral libraries (above) that contain annotated MS spectra of unique proteolytic peptides were utilized to determine modification sites specifically on Ser198-containing peptides. Raw data were input into Skyline and all MS1 peaks were manually integrated. Peak area intensities were exported as a sum of observed charge states for those peptides.

The hydrolysis time of 2.5 hours resulted in some incomplete cleavages, so to calculate the percentage of the active peptides bound to OPs, peptides with missed cleavages on both the amino and carboxyl ends of the serine active site peptides were included in calculating the normalized peak area (NPA) of active site serine residues attached to OPs. The NPA was obtained by using the (peak area of the 154 modification/ total observable peptides containing the active site including missed cleavages) *100.

## Results

### Generation of in vitro adducts with bioactivated Ops

Prior to analyzing blood samples from control and exposed individuals, candidate organophosphate esters and commonly used commercial TAP formulations were analyzed for their potential to inactivate pBChE in vitro with and without bioactivation by testing concentration-dependent inhibition of pBChE with complete bioreaction mix and complete bioreaction mix minus NADPH as well as TAP alone (**Fig. 1**). Surprisingly, significant inhibition was observed with D150 in the absence of bioactivation. This observation was interesting since D150 is the TAP blend added to one engine oil and three hydraulic fluids (**Table 1**).

**Figure 1.**
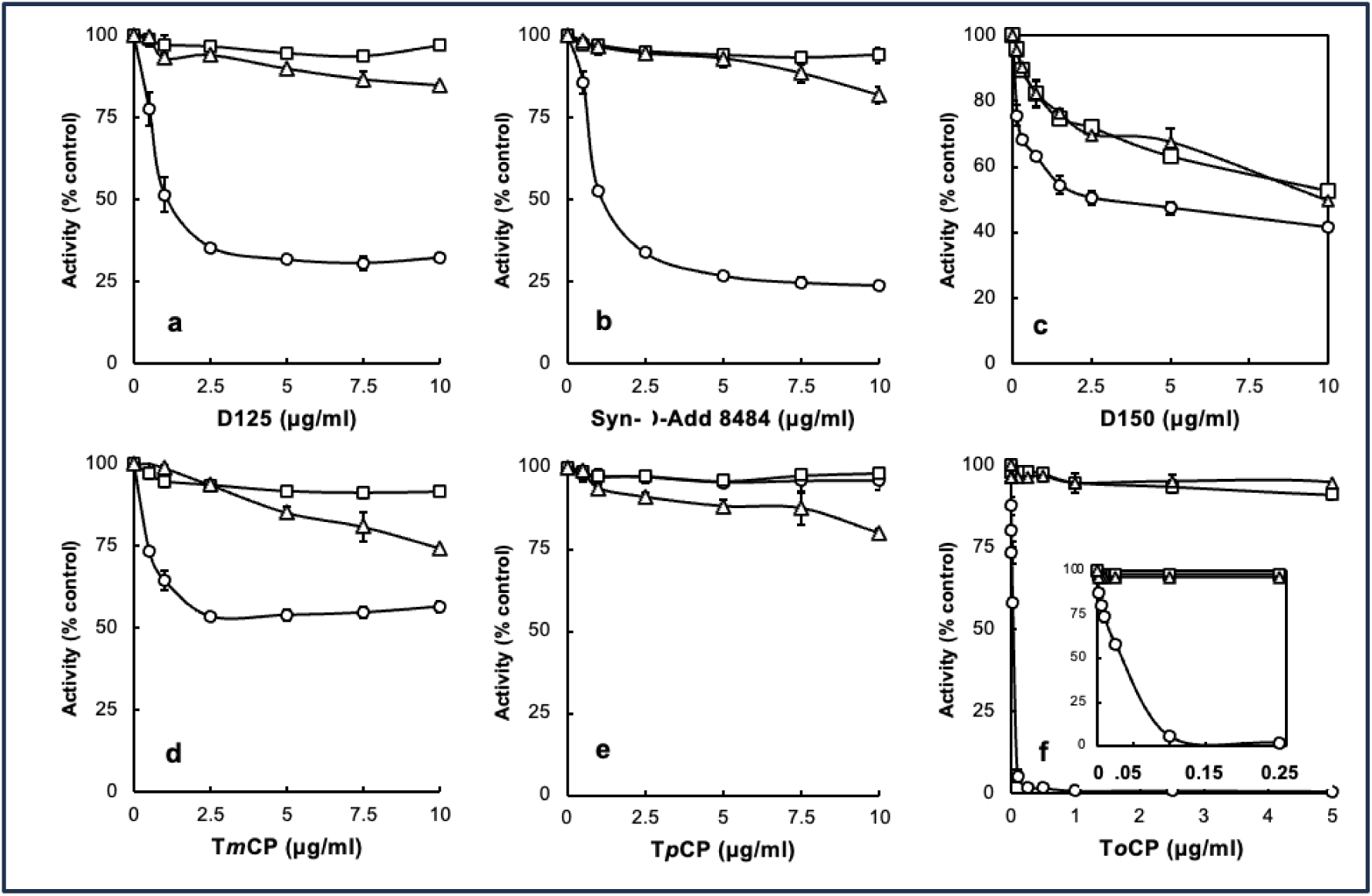
Concentration dependence of inhibition of BChE by 3 commercial mixtures of TAPs. Open circles (*O)*, complete assay; open squares (ο), complete assay minus NADPH; Open triangles (ρ) TAP only. Panel **a**, Durad 125; Panel **b**, Syn-o-Ad-8484; Panel **c**, Durad 150; Panel **d**, tri-meta cresyl phosphate T*m*CP; Panel **e**, tri-para cresyl phosphate T*p*CP; Panel **f**, tri-ortho cresyl phosphate T*o*CP (inset expanded low concentrations of T*o*CP).

**Table 1*.**
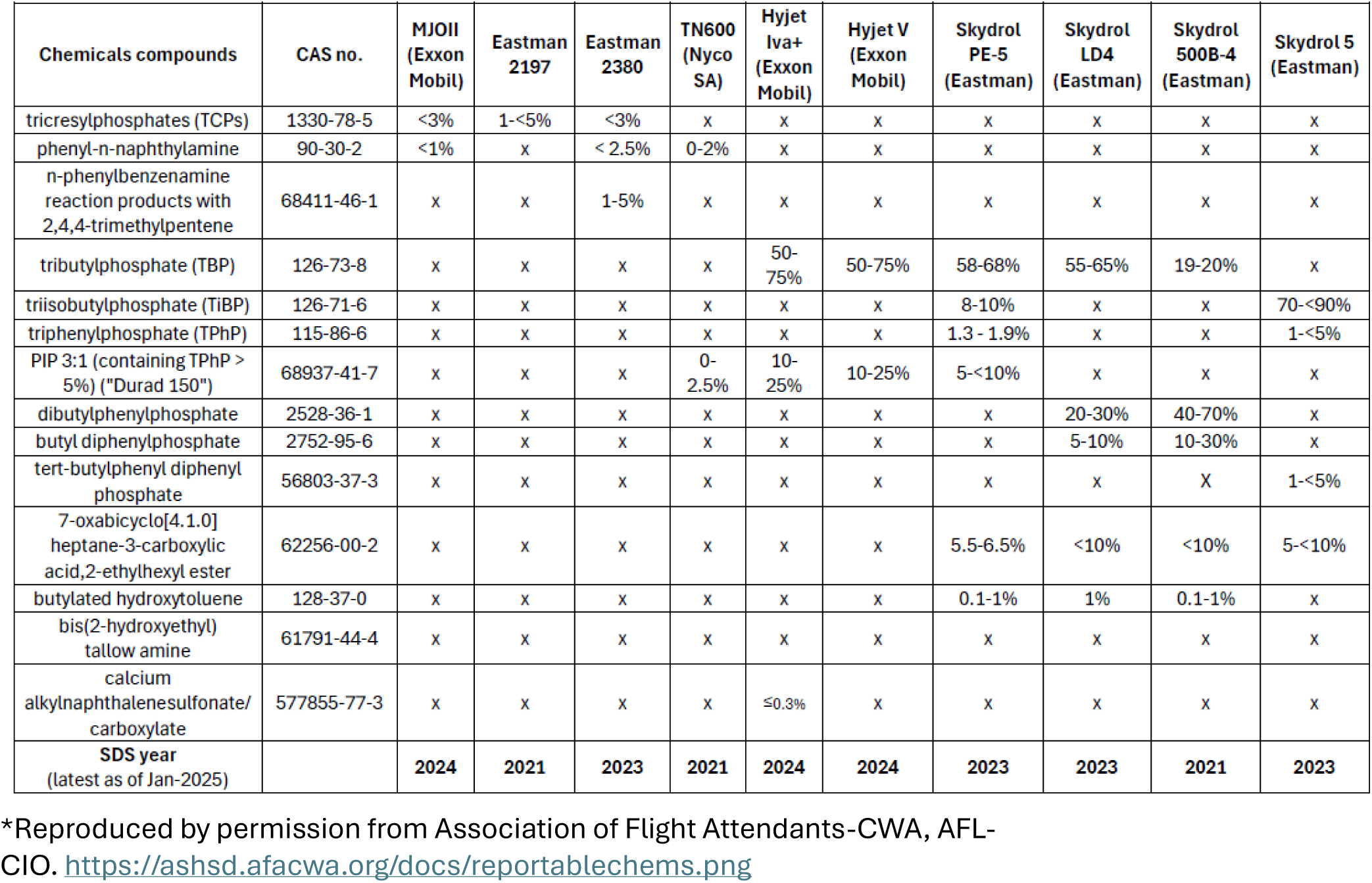
Reportable chemical compounds on a selection of aviation engine oil and hydraulic fluid safety data sheet.

Since the kinetic behaviors of D125 and Syn-O-Ad 8484® were nearly identical, we focused on D125 for LC/MS analysis of adducts. The expected and observed adducts and retention times (RT) for the tryptic peptide containing the active site serine were 1) a 186Da adduct from a cresyl phosphate with an oxidized ring methyl group (RT=19.7 min), 2) a 170Da adduct from cresyl phosphate (RT=22 min), 3) a 156Da adduct from phenyl phosphate present in the mixed ester formulations (RT=20.6 min) and 4) an 80Da phosphate adduct (RT=18.3 min) as the adducts are known to “age” to a phosphate residue (Liyasova et al. 2011). The unmodified tryptic peptide eluted at 16.8 min). Example LC/MS data are shown in **Figure 2** for the 80Da, 156Da and 170Da adducts. Points of interest in the bioactivation inhibition experiment are: 1) the inhibition of BChE by D150 that was not bioactivated; 2) the observation that bioactivated D125 and Syn-O-Ad 8484® were more effective inhibitors of pBChE than bioactivated D150 and 3) the non-bioactivated T*o*CP showed very little inhibition of BChE while the bioactivated T*o*CP was a very potent inhibitor of BChE with a Ki in the range of approximately 50 pg/mL (**Fig. 1f, inset**). This is especially interesting since Henschler (1958) reported that mono-ortho cresyl-phosphate was ten-times as toxic as T*o*CP and is present at much higher levels than T*o*CP.

**Figure 2.**
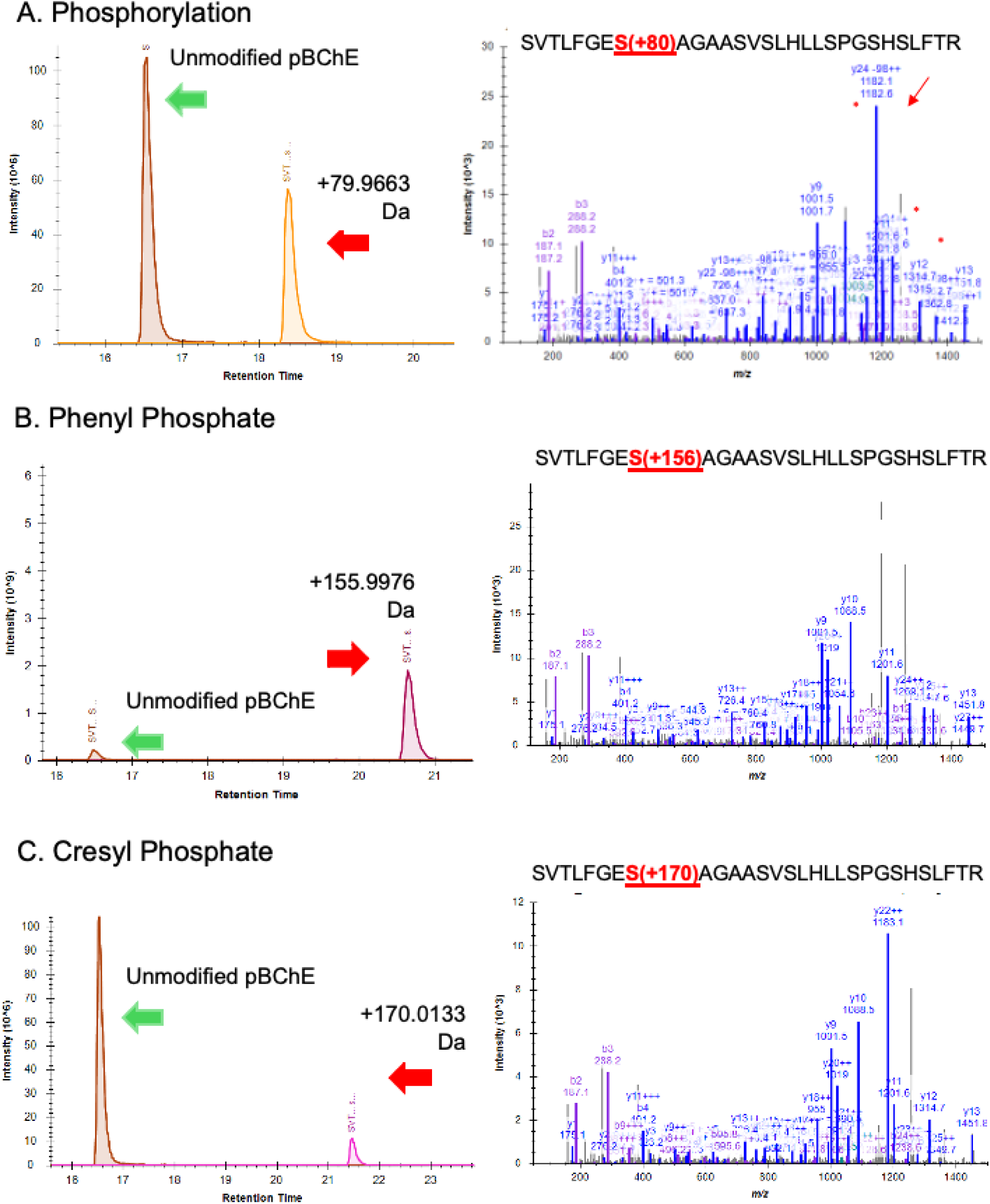
LC/MS data for the 80Da, 156 Da and 170Da adducts on pBChE generated in vitro with bioactivated D125.

### Di- and Mono-OH Me TpCPs as potential inhibitors of several potential biomarker proteins

The mono-hydroxy T*m*CP exhibited little inhibition of BChE while the Di-OH T*m*CP was an inhibitor of BChE (**Fig. 3**). In contrast, both compounds were effective inhibitors of ChE while neither exhibited inhibition of acylpeptide hydrolase from red blood cells (Kim et al. 2010). On the other hand, both mono- and di-OH T*p*CP were effective inhibitors of carboxylesterase which is discussed as one of the factors that may contribute to individuals’ different tolerances for exposure to oil fumes.

**Figure 3.**
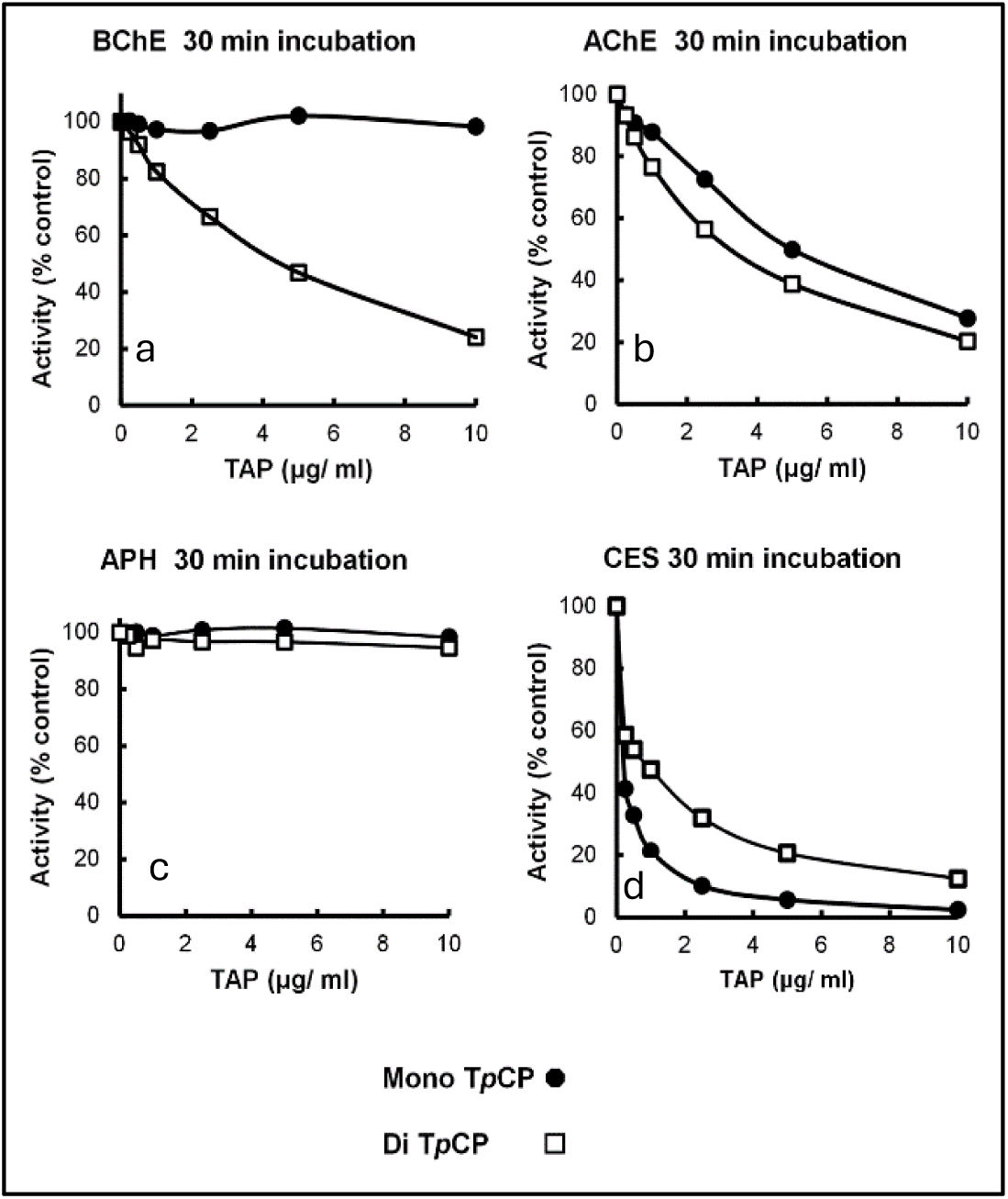
Inhibition of potential biomarker proteins by mono- or di-hydroxy para cresyl phosphate, Panel **a**, BChE; Panel **b**, acetylcholinesterase; panel **c**, red cell acylpeptide hydrolase; Panel **d**, carboxylesterase.

### Trimethylolpropane phosphate as potential inhibitor of BChE

Since TMPP is highly toxic and is formed when trimethylolpropane esters (TMPEs; the primary base stock component of one widely used aviation engine oil) react with TCPs under conditions of thermal degradation at or above 250°C which would be reasonable to expect in an operating aircraft engine (Wright, 1996). TMPP was tested as a possible inhibitor of BChE in vitro. It was not a significant inhibitor of BChE activity (data not shown) so it was not considered as a BChE adduct biomarker for exposure to OP anti-wear agents from fume events. Whether or not TMPP would inhibit BChE activity in vivo is unknown. As an inhibitor of the GABA receptor, TMPP may affect levels of downstream metabolites.

### Analysis of exposed subjects’ BChE for OP modified active site serine residues

Previously identified TCP isomer-specific adducts from *in vitro* data(e.g., 65Da, 80Da, 91Da, 107Da, 165Da, 170Da, 180Da) were not identified on the active site serine of BChE from subjects who reported onboard exposure to oil fumes. The most abundant adduct of BChE found had a mass of 154Da. **Figure 3** shows the dramatic difference in adduction of the active site serine of BChE between the majority of fume event-exposed individuals and control subjects. In addition, a commercial pool of plasma samples from ≈8,000 individuals whose flight histories were not known showed a value of 0.8% modification of the pBChE active site serine by the 154Da adduct (data not shown). Results of a typical MS analysis are shown in **Figure 4**. Control subjects were found to have similar results to that of the control pooled plasma.

**Figure 4.**
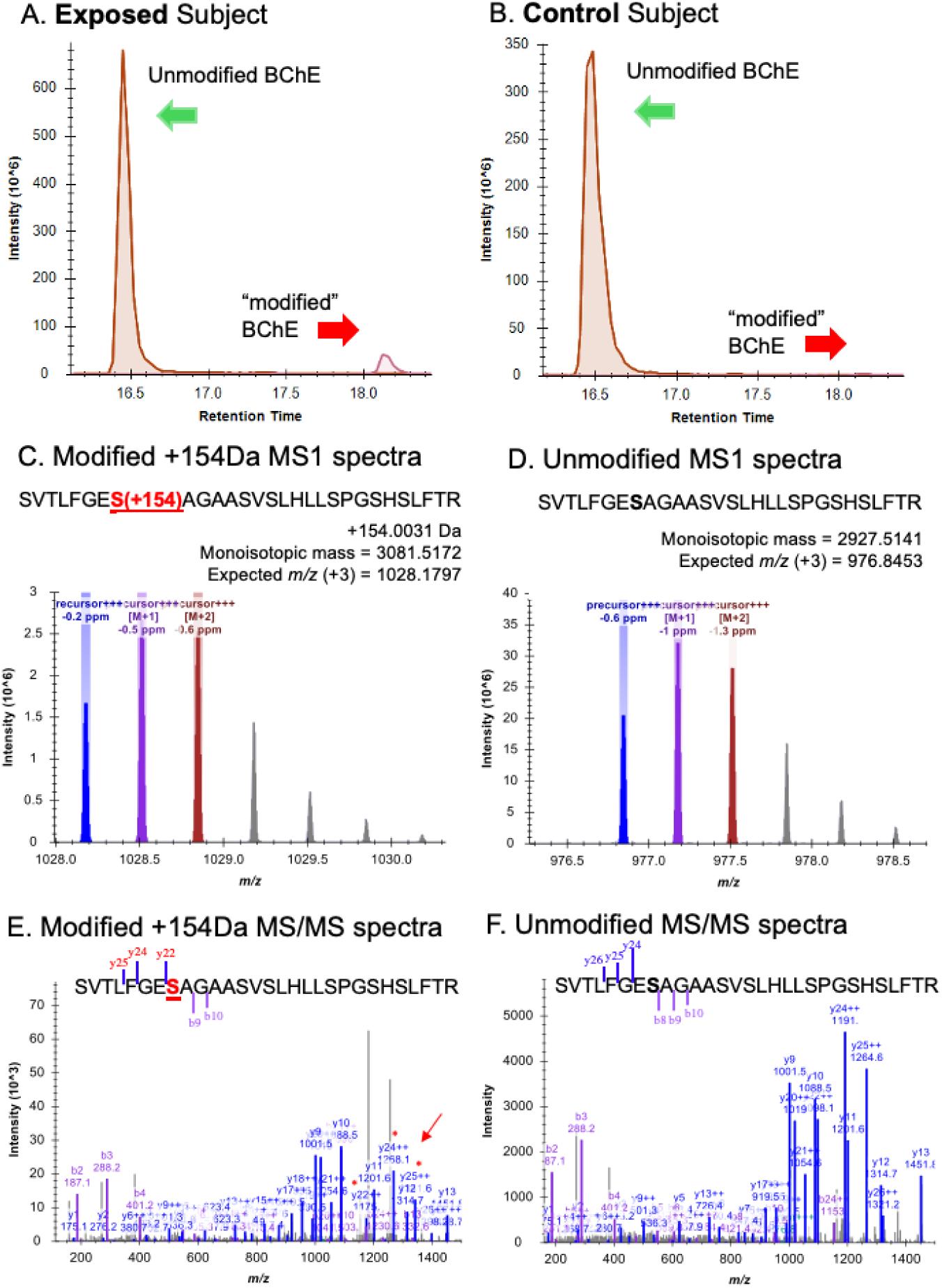
Typical analysis of the percentage modification of BChE Serine_198_ from a fume event exposed individual and an unexposed control subject.

**Figure 5.**
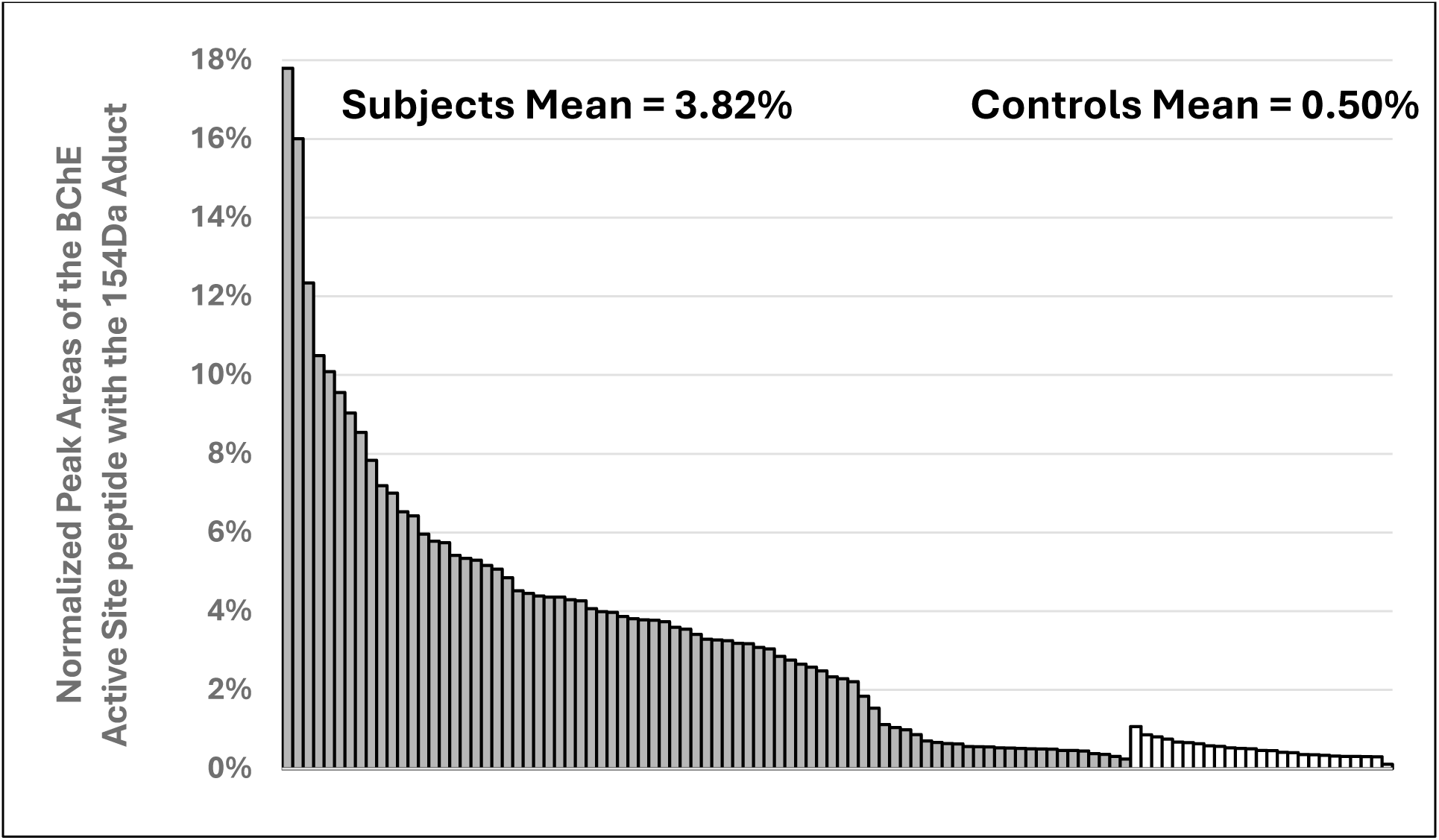
Histogram of normalized peak area of the active site peptide with the 154Da adduct on the active site serine of BChE from individuals exposed to a fume event and control subjects. Solid bars, 81 exposed subjects (37 crew, 18 passengers, 26 pilots) and open bars, 25 unexposed controls.

Examination of tables of phosphate adducts suggest that the observed adduct mass of 154.0031Da is compatible with addition of a glycerol phosphate to the BChE active site serine 198. Of 81 subjects, many samples with a normalized peak area for the 154Da adduct greater than control subjects (*X̅*=0.50%; range=0.11%-1.07%) were from fume event exposures that pre-dated 2013 (N=59); range = 0.46%-17.8%, *with X̅*=4.84% (Fig. 6). Samples from the remaining 16 subjects with fume event exposures from 2016-2024 only showed the 154Da adduct at background levels (0.24%-1.13%; *X̅*=0.55%), as confirmed in control samples from people who had not flown in at least three months. To examine the possibility that a similar mass shift could have been associated with an adduct formed with PMSF (a protease inhibitor), due to treatment of the plasma pBChE standard during purification was ruled out since none of our subject samples were treated with PMSF nor did we detect any change in the percent modification or shift in mass with in vitro experiments that directly tested for modification by PMSF. A reasonable assumption based on the existing data is that an upstream inhibition of a metabolic pathway of lipid or carbohydrate metabolism may lead to the increased level of the 154DA adduct on the BChE active site serine. Finding lower levels of this adduct in control samples may also suggest that it is a normal adduct seen on BChE and may explain a physiological role of BChE.

**Figure 6.**
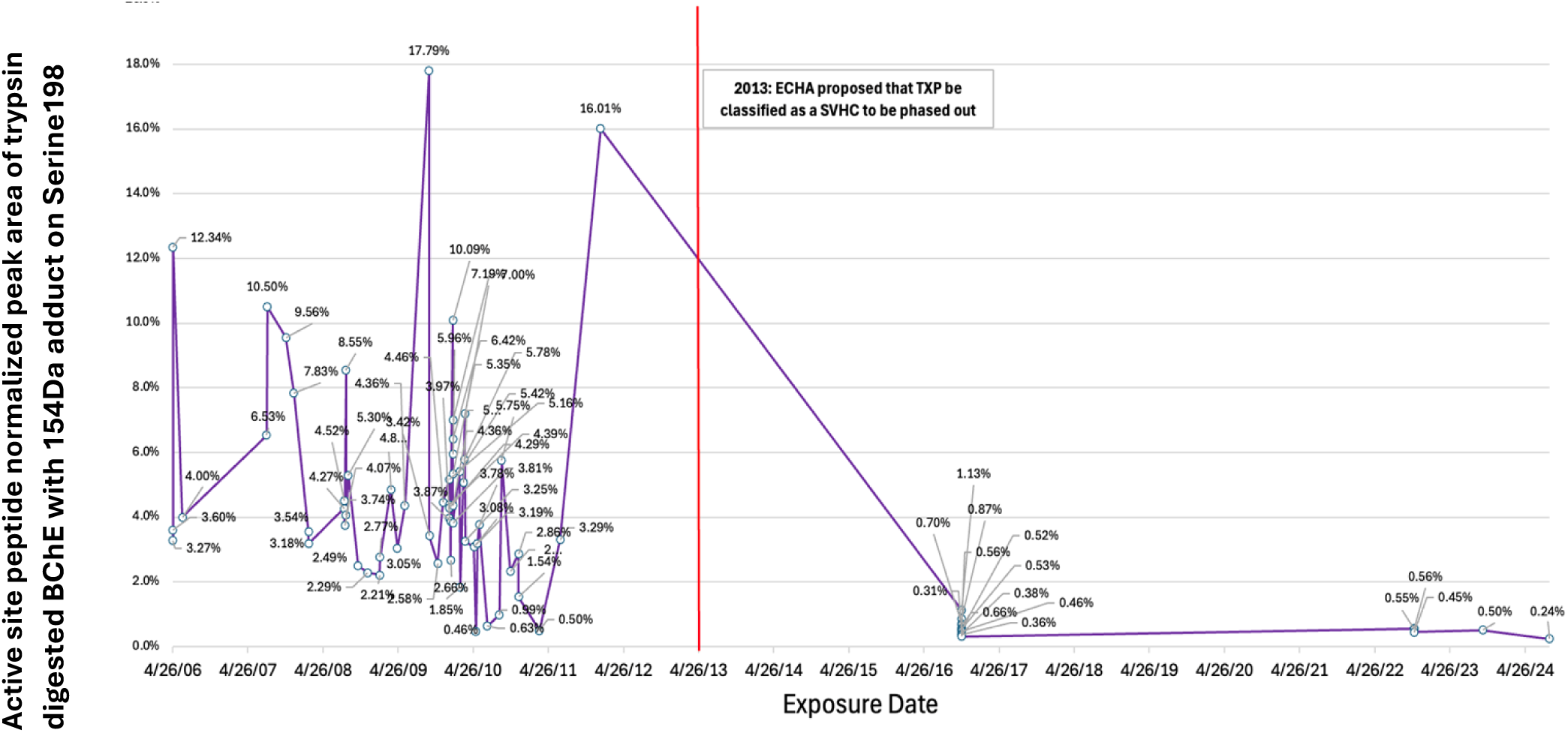
Exposure dates and normalized active site peptide peak area of trypsin digested BChE with 154Da adduct. Vertical red line, date of change to OP additive with a modified composition.

## Discussion

Due to the design of most aircraft ventilation systems, very low levels of TAPs routinely contaminate the ventilation air supplied to the cabin and flight deck (Michaelis, 2018; Johnson, 2018; Chupp et al., 2006). In addition, airlines periodically document more noticeable exposures to air supply system-sourced oil and hydraulic fluid fumes caused by various mechanical faults (e.g., worn or failed seal or bearing, spillage that gets ingested into the compressor inlet, etc.), as well as operational factors such as engine washes or overservicing an oil or hydraulic fluid reservoir (Anderson, 2021), and residues released from soiled ducting (FAA, 2025).

The normalized peak areas for the 154Da adduct were determined by MS. We also generated *in vitro* bioactivated TAPs using microsomes and the required cofactor NADPH with specific TAPs or TAP commercial mixtures and characterized the inhibition of the plasma BChE followed by MS analysis of the mass of the adduct bound to the BChE active site serine. Since there are many proteins in human blood, we made use of a rapid high efficiency immunomagnetic bead isolation of the plasma BChE (Carter et al., 2014; Marsillach et al., 2011) that provides highly purified and concentrated BChE, both from exposed subjects and controls who had not flown for at least three months.

Although modifications which inhibit BChE have been recognized as a standard indication of exposure to various neurotoxic OP compounds, oil fumes contain not one but multiple OPs. This makes exposure assessment using a simple measure of BChE activity more difficult because the sensitivity to – and extent of – inhibition can vary significantly between the different TAPs, TAP isomers, and their respective metabolites, ranging from 18% to 98% for three isomers of one TAP tested in rat serum, for example (Mackerer et al., 1999).

Another complicating factor in using BChE activity inhibition as a measure of exposure from TAPs in engine oil or hydraulic fluid leaks is that baseline BChE activity can vary significantly among individuals (La Du et al. 1990; Lockridge et al. 2016). Thus, proper interpretation of an individual’s *post-exposure* inhibition of BChE activity requires knowledge of their *baseline* BChE activity; namely, the activity determined either prior to exposure or sufficiently long enough following an exposure for the BChE activity to have returned to its baseline level. MS determination of the percentage modification of the BChE active site serine avoids the necessity of determining pre-or post-exposure basal activity individual. This is an important advantage because of the variability of basal BChE levels among individuals. Our efforts to use plasma cholinesterase active site modification methods provide useful insights into one means to explore biomarkers for OP compounds in fumes from oil or hydraulic fluid leaks.

Tricresyl phosphates (TCPs) dominate the OP blends added to engine oils. In vitro laboratory studies of the adduct mass values have identified a variety of molecular adducts from TCPs, including some that appear to apply to all isomers (91Da, 107Da, 165Da) and some that were only observed for ortho isomers (65Da, 80Da, 91Da, 107Da, 165Da, 180Da, 181Da, and 277Da) (van Netten, 2014; Rosenberger et al., 2013; Denola, 2008; Mackerer et al., 1999). None of these molecular adducts were identified on the serine active site of BChE in samples donated by the fume event exposed subjects in this study.

Instead, analysis of BChE isolated from the plasma of study subjects who reported exposure to oil fumes prior to 2012 – most of whom suffered typical symptoms of TAP exposures – showed a significant increase (p=0.00005) in a 154Da adduct on the active site serine compared to the BChE adduct data for control samples collected in 2023-24 from individuals who had not flown in at least three calendar months and had no history of documented fume events. The mean background 154Da adduct level of recent control samples (NPA - 0.5%) data reflected background level of the 154Da adduct found in a pool of plasma samples (N≈8,000). The increased NPA of the 154Da adduct in the exposed individuals from the early fume events (prior to 2012) was unexpected and not observed among the OP metabolite adducts in any of the *in vitro* exposure experiments. The 154Da adduct has a mass consistent with glycerol phosphate suggesting that it may derive from perturbation of phospholipid or carbohydrate metabolism, by *in vivo* disruption of a metabolic pathway, by a bioactivated TAP or by a catalyzed ester exchange reaction (e.g., Chamot-Rooke et al., 2011). These results also suggest a possible physiological function of BChE.

One explanation for this difference in the observed OP metabolites on the active serine site of BChE in exposed individuals compared to *in vitro* exposures is that study subjects would have inhaled oil-based OP blends subjected to high temperatures (≈200°C - 650°C) in an engine compressor. Once inhaled, enzymes in the subjects’ lungs, blood, liver, and brain would biotransform the OPs to prepare them for excretion. These biological processes would not happen in a test tube, so the OP compound(s) in plasma, for example, would not be the same as the original OPs. The 154Da adduct may have resulted from a perturbation of either a lipid or carbohydrate pathway.

Much of the research on the toxicity of TCPs has focused on the effects of ingesting the tri-ortho isomer (T*o*CP) because of accidental mass poisonings which describe the inhibition of NTE after ingesting T*o*CP and the subsequent development of organophosphate-induced delayed neuropathy (OPIDN) (Travers, 1962; Hunter et al., 1944). Crewmembers exposed to engine oil fumes do not report OPIDN, and the TCP blends added to oils may not contain more than 0.2% ortho isomer content, by weight; of that small fraction, less than one-millionth will be T*o*CP, by virtue of manufacturing process and chemistry (Howard, 2020). Thus, OPIDN as a toxicological endpoint of exposure to T*o*CP is largely unrelated to the central nervous system symptoms reported after exposure to oil fumes containing TAPs because oil fumes contain nominal amounts of T*o*CP (if any) and OPIDN is not reported; thus, the two should not be equated to each other.

Inhalation toxicity data for mixed isomer TCPs and the TAP blends added to aviation engine oils have been published. These studies have reported the effects on workers (Bock et al., 1952; Bock and Bockova, 1952), animals (Lipscomb, 1995; Aldridge, 1954), and *in vitro* tests (Houtzager et al., 2017). For example, in one study, researchers subjected rats to a four-hour inhalation exposure to different blends of TAPs, including D125 and D150, which had been heated to 650°C, and then measured the rats’ neurotoxic esterase (NTE) activity. As expected, T*o*CP exposure (positive control) inhibited NTE activity more than 86%. However, exposure to mono-ortho CP would have been more relevant since not only is it present in much higher concentrations, but it has been reported to be 10 times more toxic than T*o*CP (Henschler, 1958). Despite its very low tri-ortho-isomer content, D125 inhibited NTE by 49%. Also, D150 inhibited NTE by 35%. The team concluded that “the process of vaporization [heating] is causing a change in the compound, resulting in the potential to produce neurotoxicity." (Lipscomb et al., 1995). Another explanation is that some of the meta- and para-TCPs, phenol isopropylated phosphate (the primary constituent of Durad 150), the mono- and di-ortho isomers of TCP (present at orders of magnitude higher than T*o*CP), and other unnamed TAPs inhibited NTE. Of importance, NTE is only one measure of neurotoxicity, and may not be relevant to the central nervous system effects reported by affected airline crews. For example, trimethylolpropane phosphate is formed when TCPs are heated with the trimethylolpropane esters present in the lubricants forming a potent inhibitor of the GABA receptor with a toxicity similar to sarin (Nachon et al., 2013).

### Challenges of developing biomarkers for TAPs in oil fumes

In addition to the advantages of measuring OP-specific adducts to the active site serine of BChE, the method development work illustrates the challenges of attempting to develop biomarkers for these complex mixtures of OPs in engine oil fumes in the following ways:

### Variability in exposure chemistry

Organophosphate blends can vary by engine oil product and are different again in hydraulic fluids (Table 1) (AFA, 2025). In addition, the composition of oil and hydraulic fluid products changes over time. Table 2 illustrates the changes in the composition of reportable additives from 1988-2024 in one widely used aviation engine oil, based on what is reported on product safety data sheets (SDS).

**Table 2:**
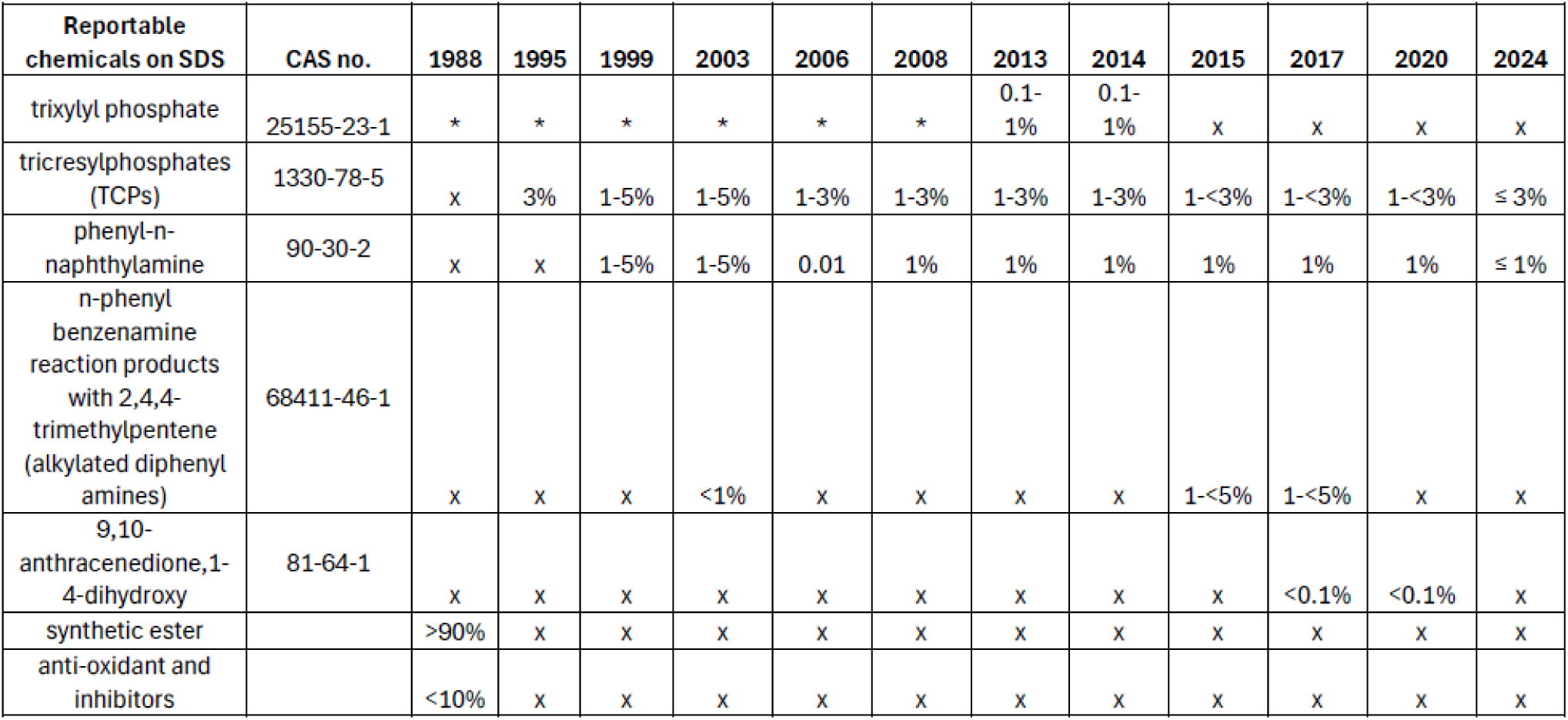
Reportable chemical compounds on Mobil Jet Oil II Safety Data Sheets, reported 1988-2024.

One documented change in TAP blends added to most engine oils was the phase out of trixylyl phosphates (TXPs; CAS no. 25155-23-1). This change was in response to stricter labeling and classification rules proposed in 2013 (ECHA, 2013) after TXP was formally characterized as a reproductive toxin. The effective ban was fully implemented in 2015. Based on an analysis of product SDS, reported TXP content changed from approximately 0.6% (2005) to less than 0.1% (2015).

Other smaller and unreported changes to the OP blends in oils may have occurred during the time frame over which blood samples were donated for this research, also, although the maximum allowed ortho isomer content of TCPs (0.2%) has not changed since 2008. Characterization of the 10-times more toxic mono-ortho isomer (Henschler, 1958) which dominates the ortho content of the TCPs in engine oils (Howard, 2020) does not seem to have been considered or tested.

It is important to note that when the term “TCP “is used on an engine oil product SDS, it is understood to mean a TAP blend dominated by TCPs. For example, in 2000, Mobil Oil toxicologists described their understanding of the usage of this term:

> *“The TCP used in jet engine oil is a very complex mixture. The conventional TCP used in Mobil Jet II is a complex mixture prepared primarily from m and p cresol. However, other substituted phenols as well as xylenols are present in the synthesis mixture. We have identified 10 phenols and xylenols, as well as low levels of ortho cresol and phenol, in hydrolyzed conventional TCP. Ortho cresol was present at about 0.16%, m + p cresol combined at 80% and the other phenols at 17%. Thus, the number of triaryl phosphate combinations in TCP is very high and is not limited to the ten that can be formed from the meta and para cresol.” (Mackerer and Ladov, 2000).*

TCP production processes may have been refined over the past 25 years such that the “other phenols” OP component in the TAP blends added to engine oils may be less than 17%. Still, the process by which TCPs are produced is not expected to generate only the collection of isomers that are represented by CAS number 1330-78-5.

Some of the volunteer subjects in this study may have been exposed to hydraulic fluid; some to a TCP-free oil used by at least two commercial airlines (Turbonycoil 600), and some to fumes that were generated from oil and other residues on the air conditioning packs or ducting which may introduce some variability to the chemical composition of onboard exposures.

Finally, aviation engine oil fumes are comprised of a complex mixture of compounds which includes – but is not limited to – OPs. For example, after heating a sample of engine oil to 350°C, researchers counted 634 distinct peaks in the gas chromatogram, only 170 of which could be identified by name (Houtzager et al., 2017). Interactions between these constituents – in the test tube and in the human body – are not well understood. Decomposition and toxicity of engine oil both increase with temperature (Lipscomb et al., 1995; Treon et al., 1955). Exposing oil to high temperatures in the engine and APU will also generate ultrafine particles (UFP) (FAA, 2023; ASHRAE. 2022). It has been suggested that UFP may act as a vehicle to transport the OPs in oil and hydraulic fumes across the blood-brain barrier (Howard et al., 2018).

The engine oils and hydraulic fluids to which subjects were exposed will have been subjected to a range of temperatures, depending if they originated in the APU compressor (lower temperature or a main engine compressor (higher temperature), and also depending on the size and power of the engine. The temperature range within which these fluids are heated is unlikely to meaningfully impact the exposure chemistry of the organophosphates (which are used, in part, for their heat stability) but will alter the chemistry of other chemical constituents and decomposition products in the fumes.

### Time lag between exposure and blood draw

The half-life of BChE offers both an advantage and a challenge. For some biomarker applications (such as urinary measurements) a biological sample must be provided within a short time of exposure (e.g., 24 hours). The longer half-life of BChE (approximately 12 days) provides additional time for sample collection. The reported time lag between exposure to oil fumes and blood draw for 73 of the subjects in this study ranged from 0.5 to 60 days with a median of seven days. The remaining 6 subjects either did not report the time lag or it was reported to exceed 60 days; also, information on air travel (if any) between the reported fume event and the date of the blood draw was not provided.

### Variability in individuals being exposed

In addition to the variability in fluid composition and environmental conditions, there are inter-individual differences in susceptibility because of the genetic differences of individuals who inhale these compounds in contaminated aircraft ventilation supply air. All these factors are expected to affect BChE concentration and activity in their blood and will also influence an individual’s tolerance to exposure and susceptibility to ill effects. These factors extend to the variability in levels of other serine active site enzyme ‘OP sponges’ and previous exposures decreasing the availability of the protective OP “sponges.” One excellent example of the variability noted in this study where a husband and wife were sitting next to each other (2008) and each showed almost identical increases in the modification of the 154Da adduct to their BChE active site serine. However, the wife had serious life-long tremors following her exposure while the husband was asymptomatic. The likely explanation is that the husband had higher levels of serine active site enzymes that bound OPs and was thereby protected by higher levels of serine active site enzymes that precipitated the pathology in his wife. Studies with the prophylactic administration of human BChE to Göttingen minipigs demonstrated protection against lethal exposures to the nerve agent sarin (Saxena et al., 2015). While the increases in levels of adduction of the active site serine with the 154Da adduct clearly correlate with the fume exposures they do not provide direct evidence of exposure to specific OPs.

At least 3 three factors form the basis of individual differences in resistance to the effects of exposures to fume events:

First, there is inter-individual variability in the levels and activity of front-line defenses (e.g., BChE, CES and other serine active site esterases, proteases, lipases, and other target proteins). Also, previous exposures will deplete or inactivate these “molecular sponges,” rendering an individual more susceptible to the effects of a future exposure if it occurs before their baseline resistance was restored, which will depend on the half-life of each protective protein.
Second, genetic differences in the levels of key enzymes involved in detoxification, binding and metabolism of TAPs are expected to influence individual susceptibility to ill effects of exposure to these compounds. For example, clinical effects of genetic differences in both BChE activity have long been recognized (La Du et al., 1991; Lockridge et al., 2016). Similarly, carboxylesterases (CESs) play a significant role in catalytically detoxifying some xenobiotics and are high affinity stoichiometric sponges for OPs. In a sample of only 12 people, the activity of 10 CES enzymes in the liver varied between five and 45-fold. (Hosokawa et al et al., 1995).
Third, some individuals are less tolerant to the chemical constituents in oil fumes than others which relates to differences in how sensitive certain serine active site enzymes are to different metabolites of TAPs. BChE, for example, not only varies in concentration among individuals by about three-fold, but genetic polymorphisms also contribute to differential sensitivity to certain OP exposures (La Du et al., 1991; Lockridge et al. 2016; Loewenstein-Lichtenstein et al., 1995). Many of the serine active site enzymes are stoichiometric scavengers, i.e., each protein molecule binds only a single OP molecule with high affinity. Nonetheless, they bind the OP with significant affinity to protect the individual against the effects of exposure to certain OPs.

### Practical application of our findings

Except for the Boeing 787 (B787) aircraft, crew and passengers on all commercial, cargo, and military aircraft may be exposed to engine oil fumes during otherwise normal flights. In the US commercial fleet, for example, an average of 5.3 fume events were documented each day over a six-year period (Shehadi et al., 2015). Crew members have a higher risk of exposure to these fumes (low-level and higher-level) than even the most frequent flyers because of the significant time that crews spend in the aircraft environment. Pilots will generally have higher levels of exposure than individuals in the cabin because the flight deck receives a higher flow rate of ventilation air compared with the cabin to manage the higher heat load and prevent any cabin smoke from entering the flight deck (Hocking, 2000). In addition, chronic low-level exposure to these TAP compounds will make crews more susceptible to the ill effects of a subsequent higher-level exposure (Howard, 2020). Given these occupational exposure scenarios, a biomarker of exposure is needed.

With respect to possible treatments for exposure to these OP blends and metabolites, it is well known that a component in grapefruit, naringenin, is a potent inhibitor of cytochrome P450 3A4 which is involved in the bioactivation of TAPs (Baker et al, 2013); however, what is not known is whether it is beneficial or not to inhibit the cytochrome P450 bioactivation because cytochrome P450 3A4 may metabolize some of the TAPs in oil fumes into metabolites that are more easily secreted. However, studies need to be conducted to answer this question, which deserves further investigation. The very high toxicity of bioactivated T*o*CP (Fig. 1f) (which has been described as up to 10-times less toxic than the mono- and di-ortho isomers; Henschler, 1958) is interesting in this respect. It should also be mentioned here that an individual’s level and activity is affected by diet, exposures as well as polymorphisms (Zanger and Schwab, 2013).

### Next steps

Here, we identify a biomarker that was consistently elevated in crewmembers with self-reported onboard exposure to engine oil fumes up to 2012. That biomarker was not elevated in the albeit smaller dataset of blood samples collected from oil-exposed crewmembers starting in 2016. This difference is likely explained by a change in the composition of the OP additives in oils across the time period of this study. The next step will be to apply these and other methods to identify one or more biomarkers associated with current engine oil fume exposures. Also, it is important to modify the sample collection and analytical methods to facilitate the use of blood spots from finger pricks which can be collected easily and quickly post-exposure by subjects. Dried blood spots can be shipped through the regular postal services, saving the inconvenience and significant expense and time associated with transporting chilled samples to an overnight shipping service, as well as organizing someone at the receiving end to process the samples on arrival. These improvements would also eliminate the difficulty and expense of locating a laboratory to draw venous blood samples. Another advantage of using the dried blood samples is that they are amenable to high through put sample analysis using either the commercial “Kingfisher” mag bead processing system available in the MS laboratory, or our laboratory constructed mag bead purification protocol in combination with a vacuum filter processing system that accommodates the use of 96 well vacuum filter plates. We have had experience with the fingerstick/dried blood spot protocol in analyzing exposure to organophosphate insecticides in agricultural workers and processors.

All in all, transitioning to blood spot collection and analysis would save significant time and expenses associated with the currently used venous blood draws, and overnight shipping of the chilled blood samples to the receiving laboratory, and would improve the sample processing at the receiving laboratories. The protocols described above would benefit from these improvements, as would the subjects exposed to fumes.

### Conclusions

An advantage of this MS-based protocol for identifying exposure to a particular OP (parent compound or metabolite) is that a baseline (or “pre-exposure”) blood sample is unnecessary, making it a useful exposure tool for workplace exposures to OPs, particularly given the significant variability in BChE concentration and activity among individuals.

Some of the challenges of using this MS-based protocol to assess exposure to oil fumes are the variability and complexity of the exposure (i.e., differences in the OP formulations, both between products and within a product over time), the exposure conditions (i.e., temperature in engine compressor and whether exposures are low-level frequent, higher-level less-frequent, or both), and the differences in the genetic and biochemical make up of subjects exposed to these fumes.

Most subjects who donated a blood sample after self-reporting a fume event onboard commercial flights showed above-background levels of a 154Da protein adduct on the active site serine of their plasma cholinesterase. This finding was not universal, likely due to variability in both the exposure chemistry and in inter-individuals’ biochemistry and genetics. Also, the elevated 154Da adduct finding appeared to be limited to blood samples collected prior to 2012. A change in the OP formulations added to engine oils over time may explain the observed time trend, highlighting the importance of further investigation into adducts on other blood proteins that more directly correlate with exposure to oil and hydraulic fluid fumes on aircraft.

Given the reported health impacts and the observed increase in the 154Da BChE adduct in the majority of exposed subjects, the best solution should be to eliminate these exposure hazards by designing future aircraft that employ dedicated electrical compressors for ventilation, such as used in the Boeing 787. Interim measures that should reduce these exposure hazards include less toxic engine oil and hydraulic fluid formulations, suitable bleed air filtration, and modifications to the existing designs, where possible. The approaches developed in this study for monitoring exposures may also be tailored for other applications.

## Data Availability

All data produced in the present study are avaialable upon reasonable request to the authors.

## Abbreviations

BChE: butyrylcholinesterase/plasma cholinesterase;
MS: mass spectrometry;
CBDP: 2-(o-cresyl)-4H-1,3,2-benzodioxaphosphoran-2-one;
D125: Durad 125;
D150: Durad 150;
DTNB: 5,5’-dithio-bis-nitrobenzoic acid;
dd-H_2_O: double-distilled water;
ECHA: European Chemicals Agency;
NADPH: nicotinamide adenine dinucleotide phosphate, reduced;
OP: organophosphate;
OPIDN: organophosphate-induced delayed neuropathy;
SDS: safety data sheet;
SVHC: substance of very high concern;
TAP: triaryl phosphate;
TCP: tri-cresyl phosphate;
HLMs: human liver microsomes;
T*m*CP: tri-meta-cresyl phosphate;
T*p*CP: tri-para-cresyl phosphate;
T*o*CP: tri-ortho-cresyl phosphate;
TMPP: trimethylolpropane phosphate;
TMPE: tri metholylpropane ester;
TXP: trixylyl phosphate.

## Acknowledgements

We are indebted to the control subjects and oil fume exposed individuals who donated blood to make this research possible. The initial research support was provided by NIEHS (Grant number P42 ES04696) and from the Royal Australian Air Force. More recent funding was provided by cabin crew unions (Association of Flight Attendants-CWA, AFL-CIO; Association of Professional Flight Attendants; British Airlines Stewards and Stewardesses Association, Canadian Union of Public Employees-Airline Division; Canadian Flight Attendant Union (CFAU); CUPE Local 4084; International Association of Machinists & Aerospace Workers District Lodge 142, AFL-CIO, Transport Workers Union, AFL-CIO; Unabhängige Flugbegleiter Organisation, Unite the Union); pilot unions (Aeropers, Air Canada Pilots’ Association, Allied Pilots’ Association, Australian Federation of Airline Pilots, Australian and International Pilots’ Association, Austrian Cockpit Association, Independent Pilots’ Association, Japan Federation of Aviation Industry Unions, Pilotenverband Swiss Ewiges Wegli, Syndicat National des Pilotes de Ligne, US Airline Pilots’ Association, Vereiningung Cockpit), other associations (Australian Licensed Aircraft Engineers Association, Clean Up Cabin Air, Global Cabin Air Quality Executive, International Transport Workers’ Federation, Sammenslutningen av Fagorganiserte i Energisektoren, Transport and General Workers Union), and individual aircrew.

